# Accumulation of meningeal lymphocytes, but not myeloid cells, correlates with subpial cortical demyelination and white matter lesion activity in progressive MS patients

**DOI:** 10.1101/2021.12.20.21268104

**Authors:** Shanzeh M. Ahmed, Nina Fransen, Hanane Touil, Iliana Michailidou, Inge Huitinga, Jennifer L. Gommerman, Amit Bar-Or, Valeria Ramaglia

**Author notes:** **Corresponding authors:** Valeria Ramaglia, Department of Immunology, University of Toronto, Toronto, Ontario M5S 1A8, Canada. and Amit Bar-Or, Center for Neuroinflammation and Experimental Therapeutics and the Department of Neurology, Perelman School of Medicine, University of Pennsylvania, Philadelphia, PA, USA, 19104.

## Abstract

Subpial cortical demyelination is an important component of multiple sclerosis (MS) pathology contributing to disease progression, yet mechanism(s) underlying its development remain unclear. Compartmentalized inflammation involving the meninges may drive this type of injury. Given recent findings identifying substantial white matter (WM) lesion activity in patients with progressive MS, elucidating whether and how WM lesional activity relates to meningeal inflammation and subpial cortical injury is of interest. Using post-mortem formalin-fixed paraffin-embedded tissue blocks (range, 5-72 blocks; median, 30 blocks) for each of 27 progressive MS patients, we assessed the relationship between meningeal inflammation, the extent of subpial cortical demyelination, and the state of subcortical WM lesional activity. Meningeal accumulations of T cells and B cells, but not myeloid cells, were spatially adjacent to subpial cortical lesions and greater immune-cell accumulation was associated with higher subpial lesion numbers. Patients with a higher extent of meningeal inflammation harboured a greater proportion of active and mixed (active-inactive) WM lesions, and an overall lower proportion of inactive and remyelinated WM lesions. Our findings support the involvement of meningeal lymphocytes in subpial cortical injury, and also point to a potential link between inflammatory subpial cortical demyelination and pathological mechanisms occurring in the subcortical white matter.

## Introduction

Subpial (type III) cortical lesions have emerged as an important component of multiple sclerosis (MS) pathology (1-7). Extensive subpial cortical demyelination has been reported in earlier stages of MS (4, 5, 8) and appears to reach maximal burden in the latter progressive stage of the disease (1-3, 6, 7). Subpial changes are not limited to demyelination but also include neuronal (9, 10) and axonal (11) damage. Importantly, the extent of cortical injury is now considered to be a major contributor to disease progression in MS including both physical (12) and cognitive (13, 14) impairments. Understanding the mechanism(s) driving subpial cortical pathology is critical to guiding development of therapies for progressive MS.

The absence of major blood brain barrier (BBB) disturbances (15), a paucity of parenchymal immune cell infiltration (1, 2), and the lack of significant terminal complement activation (16) within subpial cortical demyelinating lesions has suggested that this form of cortical injury either proceeds independently of inflammation or involves inflammatory mechanisms that differ substantially from those underlying classic deep white matter (WM) demyelinating lesions (reviewed in (17, 18)). One theory supporting a key role of immune cells in subpial cortical pathology proposes that this injury may be driven by a compartmentalized immune response that takes place within the inflamed meninges of brains that have a relatively intact BBB (4, 5, 9, 11, 19-29). While presence of meningeal inflammation has not been found to relate to either the number of white matter lesions (9) or the extent of white matter demyelination (measured as % demyelination of total white matter in whole brain coronal slices) (20), how subpial cortical injury associated with meningeal inflammation may relate to white matter lesion activity remains unknown. Such a relation is of interest, particularly in view of recent pathological findings showing that substantial lesion activity in patients with progressive MS (30). Comprehensive analysis of the autopsy cohort of the Netherlands Brain Bank (NBB) which included 3188 tissue blocks containing 7562 MS lesions from 182 brain donors with long-standing disease (>29 years), showed that 57% of all lesions were either active or mixed active/inactive and that 78% of all patients had mixed active/inactive lesions present. Notably, patients with a higher proportion of mixed active/inactive lesions at the time of death had a more severe disease course. Here we investigated the correlation between the extent of meningeal T cell, B cell and myeloid cell accumulation and associated subpial demyelination with subcortical white matter lesion activity, measured as proportion of active, mixed active/inactive, inactive and remyelinated white matter lesions derived from all available archived formalin-fixed paraffin-embedded tissue blocks (range, 5-72 blocks; median, 30 blocks) for each of 27 progressive MS patients collected at rapid autopsy from the Netherlands Brain Bank. We confirmed that meningeal immune cells are enriched in MS patients compared to controls. Stratifying MS patients into sub-groups with high versus low numbers of meningeal immune cells revealed that patients with a high number of meningeal T cells and B cells exhibited not only a higher percentage of demyelination of the subpial grey matter area, but also a greater proportion of active and mixed active-inactive lesions in the subcortical white matter, and a lower proportion of inactive and remyelinated white matter lesions. Of interest, meningeal myeloid cell numbers did not correlate with either grey or white matter pathology. While the relationship between grey matter subpial lesions and meningeal inflammation is already appreciated (9, 11, 20, 25), our data connect some aspects of white matter pathology with lymphocyte residence in the subarachnoid space in progressive MS.

## Results

### A high density of meningeal T and B lymphocytes is linked to cortical subpial demyelination in progressive MS cases

To assess the extent of meningeal inflammation and its relationship with cortical subpial demyelination, the density of meningeal lymphocytes and their topographic association with subpial demyelination were analysed in FFPE tissue blocks dissected from the supratentorial cortex of 27 MS cases compared to 9 controls. As expected, the numbers of CD3^+^ T cells and CD20^+^ B cells per unit length of the meninges were significantly increased in MS cases compared to control cases (**Fig 1a**). In addition, a significant positive correlation between the number of T cells and B cells measured in each donor per unit length of meninges was observed (**Supp Fig 1**). Considering the large dynamic range of the meningeal count for CD3^+^ T cells (range: 4.74 - 22.87 cells/mm meninges) and CD20^+^ B cells (range: 0.89 - 16.27 cells/mm meninges) in MS cases, the median was chosen as cut off to stratify the MS cases in those with high (above the median) or low (below the median) meningeal lymphocyte count (**Fig 1a** - 10.72 cells/mm for T cells; 7.30 cells/mm for B cells). Similar to other studies that have demonstrated a correlation between the presence of immune cell aggregates in the MS meninges and demyelination in the adjacent subpial brain compartment (9, 11, 20, 25), when comparing areas of subpial grey matter lesions (GML) to normal appearing grey matter (NAGM) within patients (**Supp Fig 2**), we found that T and B cell infiltrates were selectively enriched in areas of the meninges that were adjacent to subpial lesion areas compared to NAGM in the MS group with a high count of meningeal lymphocytes (**Fig 1b-g and Supp Fig 3**). It should be noted that, although increased in density, the immune cell infiltrates were never found as aggregates resembling the follicle-like structures previoulsy described (11).

**Figure 1.**
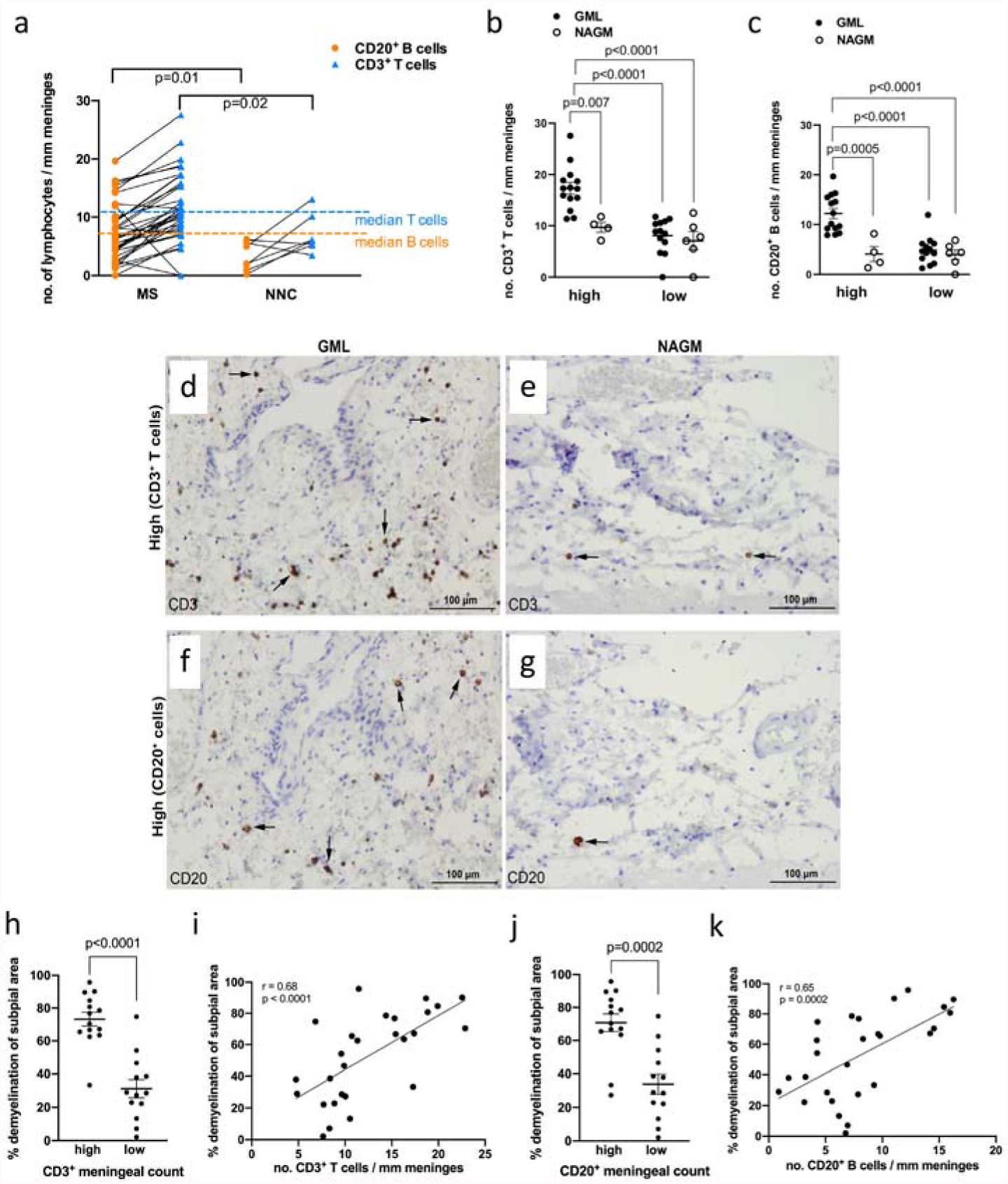
Meningeal T cells and B cells are enriched in MS and topographically linked to cortical subpial demyelination. **a**. Quantification of meningeal CD20^+^ B cell count and CD3^+^ T cell count in 27 MS donors and 9 non-neurological controls (NNC), showing enrichment in MS donors compared to NNC. The median for meningeal CD20^+^ B cell count (shown by the orange dotted line) and for meningeal CD3^+^ T cell count (shown by the blue dotted line) is used to stratify MS donors in high vs low meningeal B or T cell count. **b**. Quantification of meningeal CD3^+^ T cell count, showing a significant enrichment of T cells in meninges adjacent to GML vs NAGM in MS donors with high (n=14) but not low (n=13) meningeal T cell count. **c**. Quantification of meningeal CD20^+^ B cell count, showing a significant enrichment of B cells in meninges adjacent to GML vs NAGM in MS donors with high (n=14) but not low (n=13) meningeal T cell count. **d-g**. Representative immunohistochemical staining for CD3 (**d and e, arrows**) and CD20 (**f and g, arrows**) in meninges adjacent to a subpial grey matter lesion (GML) or adjacent to normal appearing gray matter (NAGM) in MS donors with high CD3^+^ or CD20^+^ meningeal cell count. **h**. Quantification of the percentage of demyelination of subpial area in MS donors with high versus low CD3^+^ T cell meningeal count. **i**. Spearman correlation coefficient between meningeal CD3^+^ T cell count and the percentage of demyelination of subpial area. j**h**. Quantification of the percentage of demyelination of subpial area in MS donors with high versus low CD20^+^ B cell meningeal count. **k**. Spearman correlation coefficient between meningeal CD20^+^ B cell count and the percentage of demyelination of subpial area. In **a, b, c** each data point represents the mean cell count (mean±SD) in all fields analyzed per case. Statistically significant differences were determined by the non-parametric Mann Whitney test (in **a, h, j**) or the non-parametric Kruskal-Wallis test followed by Dunn’s correction for multiple comparisons (in **b, c**). In **d-g**, scare bars represent 100μm.

Using the “high versus low” stratification for the extent of meningeal inflammation, we next determined if the density of T/B cells in the meninges was associated with clinical metadata parameters, or cortical grey matter demyelination at autopsy. With respect to the former, we found that age at the time of death and disease duration were not significanlty different for MS patients with high versus low meningeal T or B cell counts (**Supp Fig 4a-b and Table 1)**. With respect to the latter, using the entire archived collection of tissue blocks per donor, which included a comparable number of dissected blocks from donors with high vs low meningeal cell counts, we found that donors with high meningeal lymphocyte counts also had a significantly higher number of overall cortical GMLs compared to donors with low meningeal lymphocyte counts (Supp **Fig 5a, b)**. However, in terms of the types of GML, the proportion of leukocortical (Supp **Fig 5c, d)**, intracortical (Supp **Fig 5e, f)** or subpial GMLs (**Fig 5g, h**) did not differ between MS donors with ‘high’ vs ‘low’ meningeal inflammation. Since subpial cortical grey matter lesions can extend over large areas also affecting multiple gyri we next measured the percentage of demyelination of subpial grey matter in the same tissue blocks in which we assessed the meningeal lymphocyte counts. We found that donors with high meningeal lymphocyte counts also had a significantly higher percentage of subpial cortical demyelination compared to donors with low meningeal lymphocyte counts. In addition, we assessed the relationship between meningeal lymphocytes and the extent of subpial demyelination on a per-donor basis by a non-parametric *Spearman’s* correlation analysis and observed that the density of meningeal T cells and B cells positively correlated with the percentage of demyelination of the subpial area in the MS cases (**Fig 1h-k)**. In summary, while the density of meningeal T and B lymphocytes is not associated with donor clinical metadata (age of death, mean disease duration), donors with high density of meningeal T and B lymphocytes exhibit a greater extent of subpial demyelination compared to donors with low meningeal T and B lymphocytes.

### Meningeal lymphocytes are associated with white matter lesion activity in progressive MS

Although an association between meningeal inflammation and sub-pial grey matter injury has been described in multiple studies (4, 5, 9, 11, 19-29), two studies have looked at whether a relationship exists between meningeal inflammation and the number of white matter lesions (11) or the percentage of demyelination in the subcortical white matter (20) showing no link. However the relationship between meningeal inflammation and the type of lesion activity in the subcortical white matter has not been examined. We first asked whether the density of meningeal lymphocytes was associated with the proportion of active, mixed active-inactive, inactive or remyelinated WMLs obtained from the entire archived collection of tissue blocks per donor. While MS donors with high vs low meningeal T or B cell count did not differ with respect to the total number of subcortical WMLs (**Supp Fig 6a, b**), as seen in another study (11), we found that donors with high density of meningeal T or B cells had a significantly greater proportion of active (**Fig 2a, b**) and mixed active-inactive WMLs (**Fig 2c, d**). In contrast, donors with a high meningeal T or B cell count exhibited a significant lower proportion of inactive (**Fig 2e, f**) and remyelinated WMLs (**Fig 2g, h**) compared to donors with low lymphocyte meningeal count. Of note, active and mixed active-inactive lesions (unlike inactive lesions) harboured perivascular cuffing of T and B cells (**Supp Fig 7 and Supp Fig 8**), which have been linked to enriched meningeal immune cells in another cohort of MS patients (20). Altogehter, these data reveal a link between the enrichemnt of meningeal T and B cells and the WML activity in the subcortical compartment.

**Figure 2.**
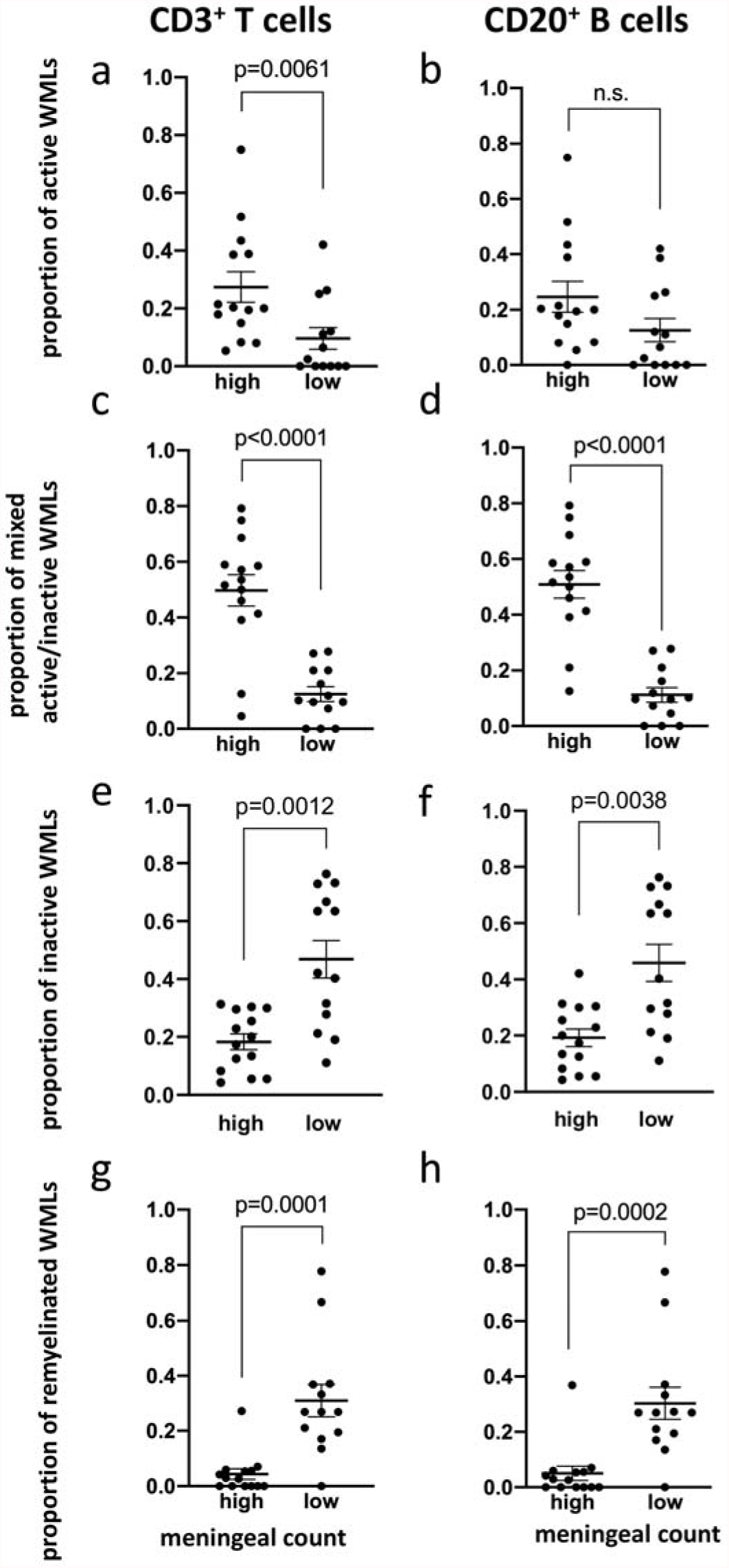
Enrichment of meningeal T cells and B cells is linked to subcortical white matter lesion activity. Quantification of (**a, b**) the proportion of active white matter lesions (WMLs), (**c, d**) mixed active-inactive WMLs, (**e, f**) inactive WMLs and (**g, h**) remyelinated WMLs in MS donors with high (n=14) vs low (n=13) meningeal CD3^+^ T cells (**a, c, e, g**) or CD20^+^ B cell (**b, d, f, h**) count. Each data point represents the proportion of WMLs in all tissue blocks analyzed per case (range 5-72 blocks, median: 30 blocks per donor). Statistically significant differences were determined by the non-parametric Mann Whitney test.

### Absence of a numerical or topographical association between meningeal myeloid cells density and subpial or white matter pathology in progressive MS

We next assessed whether similar associations between meningeal lymphocyte density and brain pathology were observed for meningeal myeloid cells by staining the same tissues with an antibody that detects ionized calcium binding adaptor molecule 1 (Iba-1) on microglia/macrophages and an antibody that detects CD68 on the lysosomal membrane of cells (31). While CD68 is expressed by a number of cell types, it is enriched on macrophages and other mononuclear phagocytes, and therefore used to identify myeloid cells, particularly those with ongoing phagocytic activity (32). Since a non-parametric *Spearman’s* correlation analysis on a per-donor basis showed that the number of CD68^+^ cells per mm length of meninges had a strong positive correlation with the number of Iba-1^+^ cells per mm length of meninges (**Supp Fig 9**), we subsequently report only on the analysis of the Iba-1^+^ myeloid cells. The density of Iba-1^+^ meningeal cells was significantly increased in MS cases compared to control cases (**Fig 3a**). Similar to the findings for T and B cells, we also found a wide dynamic range of the meningeal count of Iba-1^+^ myeloid cells (range: 11.76 – 34.64 cells/mm meninges) (CD68^+^ cells range: 17.01 – 62.57 cells/mm meninges; median 27.89 cells/mm meninges) in MS cases. Therefore, the Iba-1^+^ median (22.47 cells/mm meninges) was chosen as the cut off to stratify MS cases with high versus low myeloid meningeal density (**Fig 3a**). As we observed for lymphocytes, neither the age at the time of death nor disease duration differed in myeloid high versus low subgroups (**Supp Fig 10 and Supp Table 1**).

**Figure 3.**
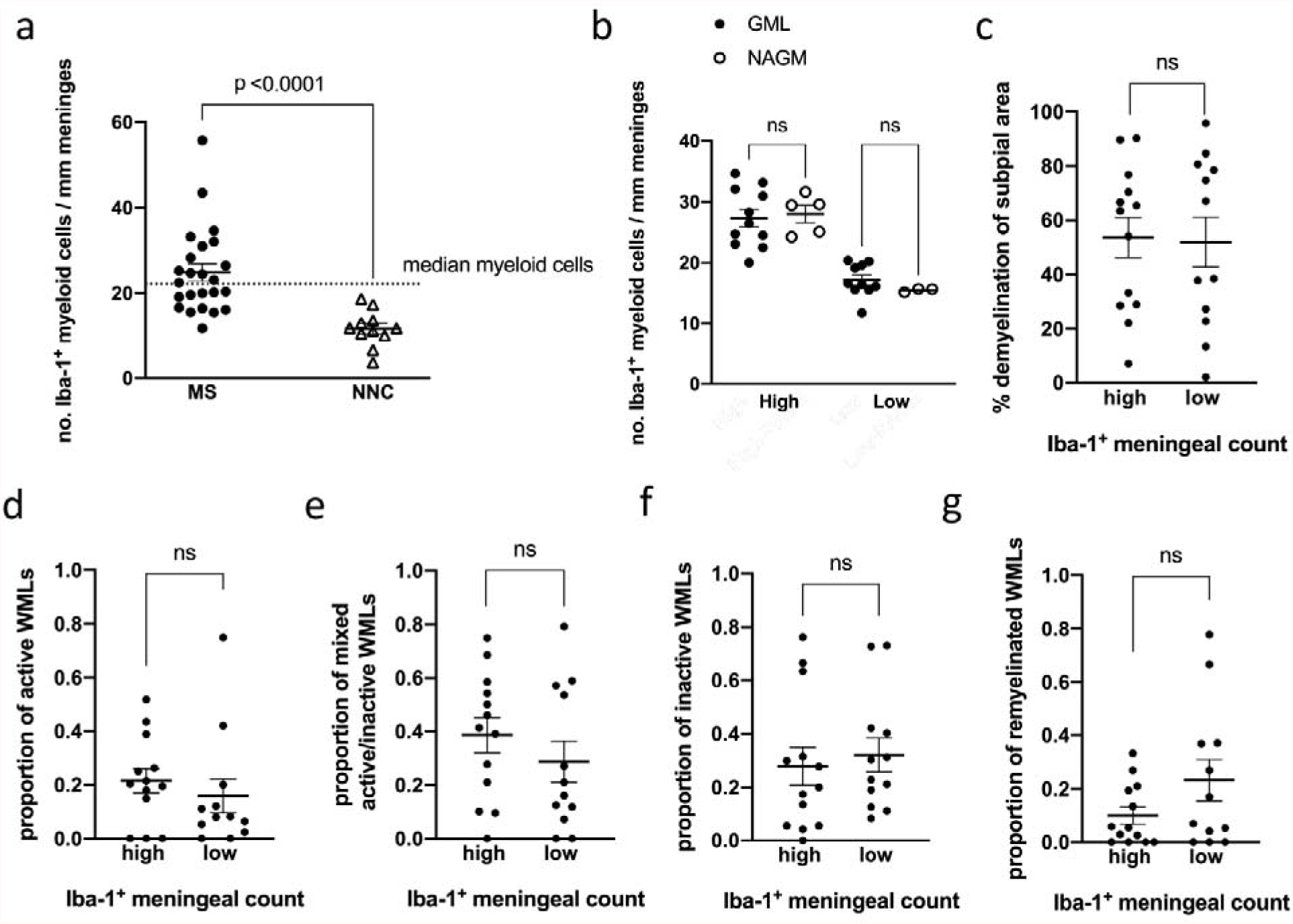
Meningeal myeloid cells are enriched in MS but not topographically linked to subpial demyelination nor to the extent of cortical subpial demyelination nor to subcortical white matter lesion activity. **a**. Quantification of meningeal Iba-1^+^ myeloid cell count in MS donors and non-neurological controls (NNC), showing enrichment in 27 MS donors compared to 9 NNC. The median for meningeal Iba-1^+^ myeloid cell count (shown by the dotted line) is used to stratify MS donors in high vs low meningeal myeloid cell count. **b**. Quantification of meningeal Iba-1^+^ myeloid cell count, showing no significant changes in the number of meningeal myeloid cells adjacent to GML vs NAGM in MS donors with high (n=14) or low (n=13) meningeal myeloid cell count. **c**. Quantification of % demyelination of subpial area in MS donors with high vs low meningeal Iba-1^+^ myeloid cell count. **d-g**. Quantification of (**d**) proportion of active white matter lesions (WMLs), (**e**) mixed active-inactive WMLs, (**f**) number of inactive WMLs and (**g**) number of remyelinated WMLs in MS donors with high vs low meningeal Iba-1^+^ myeloid cell count. Each data point represents the proportion of WMLs in all tissue blocks analyzed per case (range 5-72 blocks, median: 30 blocks per donor). In **a, c-g**, statistically significant differences were determined by the non-parametric Mann Whitney test. In **b**, statistically significant differences were determined by the non-parametric Kruskal-Wallis test followed by Dunn’s correction for multiple comparisons.

Unlike what we observed for meningeal lymphocytes, quantification of the number of myeloid cells in meningeal areas adjacent to subpial grey matter lesions (GML) or to normal appearing grey matter (NAGM) showed no spatial relation to GMLs in either groups (high versus low myeloid cell density) (**Fig 3b and Supp Fig 11**). In addition, the percentage of demyelination of subpial area did not differ between either high versus low myeloid cell density donor subgroups (**Fig 3c**). Similarly, MS donors with high vs low meningeal myeloid cell densities exhibited a similar total number of subcortical WMLs (**Supp Fig 12**), as well as comparable proportions of active (**Fig 3d**), mixed active-inactive (**Fig 3e**), inactive (**Fig 3f**) or remyelinated WMLs (**Fig 3g**). Altogether, these data show that while myeloid cells are enriched in the meninges of progressive MS patients, they are not linked to subpial cortical demyelination nor subcortical WML activity.

### The density of meningeal T and B cells directly correlates with the extent of subpial GML as well as active and mixed active-inactive WML in progressive MS

To assess the relationship between meningeal immune cells and brain pathology on a per-donor basis, we performed a non-parametric *Spearman’s* correlation analyses for MS cases. We observed that the density of meningeal T and B cells positively correlated with the proportion of active and chronic active-inactive WMLs, but correlated negatively with the number of remyelinated lesions. In contrast, we observed no correlation, positive or negative, between the density of meningeal myeloid cells versus the proportion of WMLs (**Fig 4a-l**). These data suggest a link between the density of meningeal T cells and B cells (but not myeloid cells) with subpial cortical pathology and active white matter pathology, as well as poor repair processes in WMLs.

**Figure 4.**
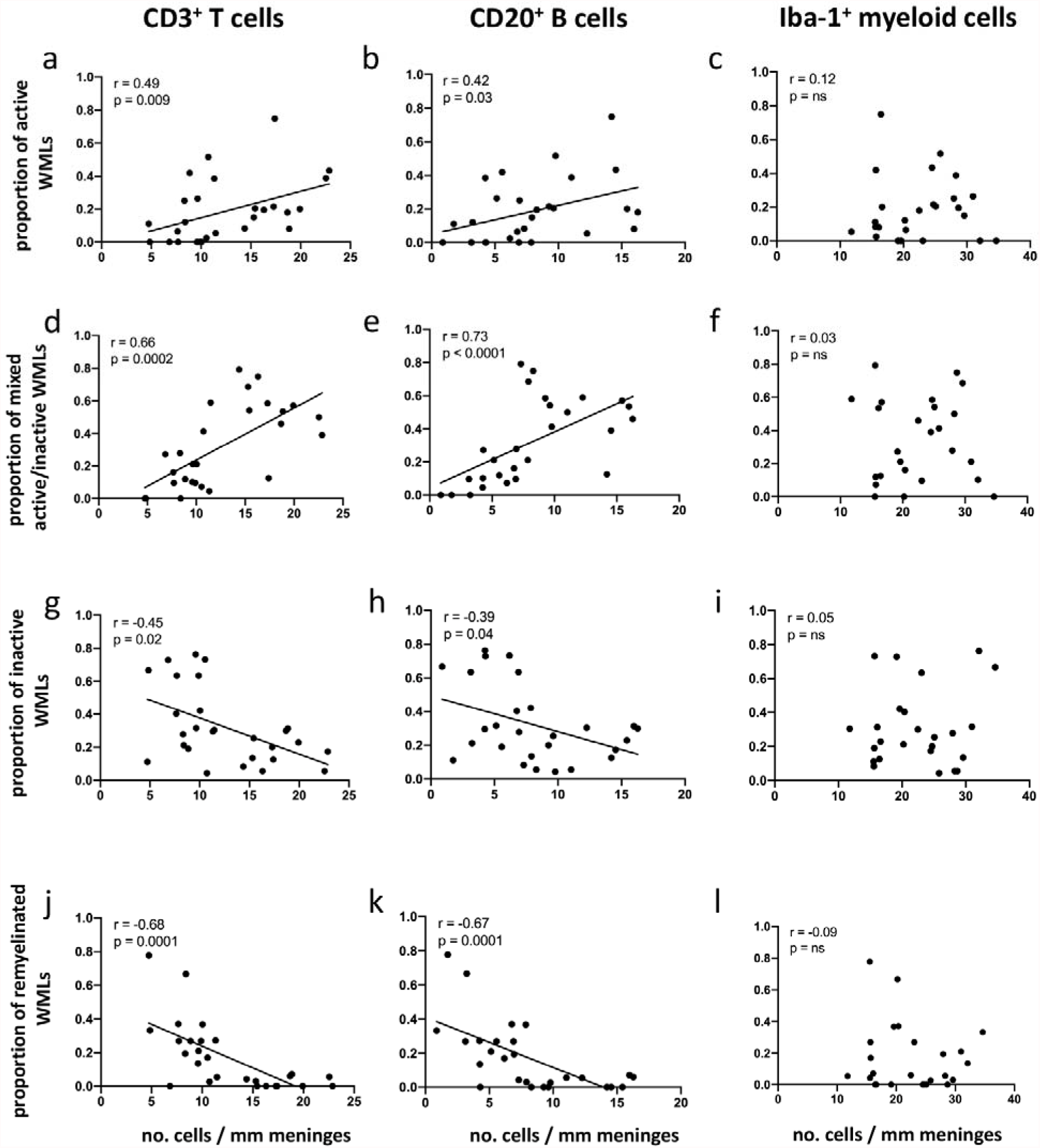
The density of meningeal CD3^+^ T cells and CD20^+^ B cells but not Iba-1^+^ myeloid cells correlates with subcortical white matter lesion activity. Spearman correlation coefficient between meningeal CD3^+^ T cell, CD20^+^ B cell and Iba-1^+^ myeloid cell count with proportion active white matter lesions (WMLs) (**a-c**), proportion of mixed active-inactive WMLs (**d-f**), proportion of inactive WMLs (**g-i**) and proportion of remyelinated WMLs (**j-l**) in each of 27 MS donors. Each data point represents the proportion of WMLs obtained from all tissue blocks analyzed per case (range 5-72 blocks, median: 30 blocks per donor) and the mean meningeal cell count in all fields analyzed per case.

## Discussion

In this study, we establish a link between meningeal lymphocytes, subpial cortical demyelination and subcortical WML activity. Specifically, progressive MS brain donors with the highest densities of B and T cells in the meninges not only exhibit greater subpial cortical demyelination (as shown previously by others (9, 11, 20, 25)) but also have greater proportions of active and mixed active-inactive lesions in their subcortical white matter. These data point to a connection between pathological mechanisms occurring in the subcortical white matter and inflammatory subpial cortical demyelination. In addition, an important implication of our findings is that pathological studies aimed at assessing potential relationships between features of meningeal inflammation and subpial cortical tissue should focus on pathologic material in which the presence of subcortical WML activity is documented.

In contrast to meningeal T cells and B cells, we noted that although myeloid cell densities in the meninges were increased in MS brains compared to control brains, myeloid cell abundance in meninges of MS patients was unchanged when comparing regions proximate to GML or NAGM. This suggests that meningeal myeloid cells are either not involved in the subpial cortical injury of MS, or that they only exert pathological activity in the presence of neighbouring T and/or B cells. Another possibility which our findings do not preclude is that while the numbers of meningeal myeloid cells do not differ between regions proximate to GML or NAGM, they may differ in their functional response profile. Lastly, since our study is necessarily cross-sectional, it is possible that myeloid cells perform important function/s in the meninges that precedes the entry of lymphocytes. Longitudinal assessment of the presence of different types of meningeal immune cells at different stages of the disease would require advanced PET imaging and MRI techniques(33) and/or the application of a relevant animal model that captures meningeal inflammation and other aspects of MS pathology, as we have described before (34, 35).

While we did not detect particularly dense immune cell aggregates (or ‘follicle-like’ structures) in the meninges of any of the patients in our progressive MS cohort, our quantitative analysis of meningeal immune cells demonstrated numbers of T cells, B cells and myeloid cells that were very much in keeping with quantification of these cells reported in prior pathological studies, which included detection of B-cell rich immune cell aggregates or diffuse meningeal inflammation in patients with progressive forms of the disease (20, 22). The absence of follicle-like structures in these tissues may reflect a technical issue – importantly, the NBB dissection protocol involves stripping of the meninges from the brain at autopsy, such that only meninges that are trapped within the sulci remain in the brain tissue-blocks. While the tissue blocks in our hands did have sufficient amount of intact meninges (about 70% of surface area analysed contained meninges) to allow for the quantification of meningeal immune cells, this preparation could explain why follicle-like meningeal structures were not detected in the tissue we examined. Another possibility for the lack of B-cell rich immune cell aggregates in our cohort could be the older age at death compared to other studies which have identified such meningeal immune cell aggregates. In our study, the age at the time of death of MS patients ranged from 41 to 81 years (median: 58 years). Magliozzi et al (11) found that meningeal immune cell aggregates could be detected only in patients with younger age at death (median: 42 years in Follicle^+^ versus median: 55 years in Follicle^-^).

Our findings of a selective association between increased numbers of meningeal T cells and B cells with the extent of subpial GML do not prove causality nor direction of causality, though they are nonetheless consistent with a potential contribution of T cell- and/or B cell-secreted factors in propagating subpial cortical injury. While we did not find a link between the extent of meningeal inflammation and the number of subpial cortical lesions, it would be relevant to investigate whether a link exists between the extent of meningeal inflammation and the activity of the subpial cortical lesions. However, we were unable to address this question in our cohort since it did not include subpial cortical lesions with evidence of activity (i.e. a rim of Iba-1^+^ or CD68^+^ cells), likely because the cohort used for the studies described here includes donors with long-standing disease. Therefore this question should be addressed in a cohort of donors with shorter disease duration. Both T cells and B cells can release factors that may be toxic to neural tissue and, for B cells, such effects may be both antibody-related or antibody-independent (36). In this regard, of interest are reports that secreted products of B cells derived from patients with MS (but not from controls) can induce apoptotic cell death in oligodendrocytes (37) as well as neurons (38), a toxicity that was neither antibody nor complement dependent.

With respect to the link between meningeal inflammation and WMLs, it is possible that classical WM focal perivascular inflammation and meningeal inflammation in MS are independent, and simply overlap temporally in brains while there is ongoing disease activity. Prior pathological studies considering such a relationship have reported that the degree of WM inflammation (defined by the distribution of MHC class II+ microglia and/or macrophages across the lesion) did not appear to differ between secondary progressive MS donors with or without meningeal cell aggregates (11). The same study, however, showed that donors with more dense meningeal immune cell aggregates did tend to have substantially more B cells and plasma cells in their perivascular WML cuffs, while brains with little meningeal inflammation exhibited only occasional B cells and plasma cells in their perivascular WML cuffs (11). Indeed, we also confirmed that active/mixed active-inactive WML unlike inactive WML did show perivascular cuffs of B and T cells. More recent studies have shown a link between inflammation in the two compartments. Fransen et al (39) reported that donors with enriched CD20^+^ B cells in the meninges overlaying the brainstem also had enriched CD20^+^ B cells in the perivascular cuffs within lesions in the brainstem. Reali et al (40) reported that the density of CD20^+^ B cells in the spinal leptomeninges correlated with the density of white matter perivascular CD3^+^ and CD20^+^ lymphocytes.

If a causal link exists between meningeal inflammation and WML, two different models may be envisioned. According to one model, destruction of myelin and axons by microglia/macrophages within active or mixed active-inactive WMLs could release myelin and axonal antigens that could potentially find their way to the draining cervical lymph nodes (perhaps via the glymphatics (41)). After priming and polarizing events occurring in the lymph nodes, activated lymphocytes more efficiently traffic into the CNS, including the meninges. It is also noteworthy that investigation of cells of the B-cell lineage (B cells, plasmablasts and plasma cells) isolated from different sub-compartments of the inflamed MS CNS (meningeal, parenchymal WM and cerebrospinal fluid) demonstrated shared B cell clones across these compartments in the same brains and suggested that the majority of the secondary diversification events (affinity maturation and class switch) are occurring within the draining cervical lymph node (42). Once in the CNS, lymphocytes can they can take up residence in the meninges perhaps via entry through the blood-meningeal barrier (43).

More recently, the discovery of the existence of functional lymphatic vessels in the meninges (44, 45) has indicated a relevant link between the CNS and peripheral immune system, perhaps providing a form of communication between these different compartments (meninges, brain parenchyma and lynph nodes). Indeed, experimental studies have shown that tracers and proteins injected into the brain parenchyma and/or the cerebrospinal fluid find their way to the cervical lymph nodes (45-47). Meningeal lymphatics seem to be key players in this communication since they assist in the drainage of cerebrospinal fluid components and meningeal immune cells, enabling immune cells to enter draining lymph nodes in a CCR7-dependent manner (48). That this communication is at play during neuroinflammation has been shown in the experimental autoimmune encephalomyelitis (EAE) model of MS where decreasing lymphatic drainage, using a Visudyne-based approach, diminished acquisition of encephalitogenic properties by antigen-specific T cells, with resulting improvement of clinical symptoms of EAE (48).

Once established in the meningeal compartments, lymphocytes could secrete by-products that are potentially inflammatory, cytotoxic and myelinotoxic. These noxious mediators can occupy the CSF circulating within the subarachnoid space from which they can cross the pial membrane and diffuse into the underlying grey matter resulting in subpial GML, as supported by studies that have correlated high cortical lesion load with proinflammatory cytokines (IFNγ, TNF, IL2, and IL22) (49, 50), molecules related to sustained B-cell activity and lymphoid-neogenesis (CXCL13, CXCL10, LTα, IL6, and IL10) (49), B-cell survival factors (BAFF), factors indicative of BBB leakage (fibrin, complement and coagulation factors) and iron-related proteins (free-hemoglobin and haptoglobin) in the CSF (28).

An alternative possibility is that damage to myelin and axons in active and mixed active-inactive WMLs could trigger retrograde degeneration propagating backwards towards cortical neurons, resulting in demyelination and neurodegeneration giving rise to GMLs. Consistent with the idea that WMLs may drive the formation of GMLs, several cross-sectional MRI studies have reported significant correlations between the total grey matter volume and the total white matter volume in T1- and T2-weighted lesions (51-54). Moreover, several longitudinal MRI studies have shown an association between the volume of white matter lesions and the loss of GM volume (55), ventricular enlargement (56) and upstream GM atrophy of the visual cortex (57). In this model, the primary formation of the GML is followed by the secondary recruitment of immune cells to the portion of the meninges that are adjacent to the GML.

In summary, our findings indicate that there is some form of communication between WML and meningeal inflammation. Future studies elucidating the significance of such a relationship between these CNS ‘sub-compartments’ will be important to better understand the mechanisms of cortical injury in progressive MS and how these may be targeted therapeutically. To this end, multiplexed tools to phenotype cells in situ in post-mortem brain tissue, as we have done before for the MS brain (58) and as others have done for COVID brains (59), are ideal to phenotype immune cells in the meninges vis a vis subpial cortical lesions and is the current focus of our ongoing studies. In addition, high throughput unbiased analysis of isolated meninges will uncover hits that can guide our understanding of the cell-type composition and pathways that are at play in the meninges of progressive MS patients with a high proportion of mixed active-inactive white matter lesions (and enriched meningeal immune cells) vs those with a high proportion of inactive white matter lesions (and a paucity of meningeal immune cells).

## Subjects and Methods

### Post-mortem tissue retrieval and inclusion criteria

Tissue blocks for this study were obtained from the Netherlands Brain Bank (NBB; Amsterdam, The Netherlands). For the characterization of meningeal immune cells, sample from twenty-seven donors with progressive (primary progressive, PP, or secondary progressive, SP) MS tissues were selected based on the presence of meninges adjacent to cortex in the tissue blocks. For the characterization of MS subcortical white matter lesions, all available archived formalin-fixed paraffin-embedded (FFPE) tissue blocks for each of 27 MS patients (range 5-72 blocks, median: 30 blocks per donor) were analyzed (see **Supp Table 1**). Tissue blocks were dissected based on the identification of lesions as guided by macroscopical examination and/or by post-mortem MRI (since 2001) of 1cm-thick coronal brain slices (30). The tissue blocks used for the analysis of meningeal inflammation and subpial demyelination performed in this study were dissected from the supratentorial cortex at locations that included the occipital or the parietal or the temporal or the frontal lobes. Donors with the myelocortical (60) variant of MS were not included in this study.

Detailed clinical-pathological and demographic data of all donors are provided in **Supp Table 1**. The age at the time of death of MS patients ranged from 41 to 81 years (median: 58 years) with a mean post-mortem delay of 8 hours and 31 minutes (SD, ± 1 hour 42 minutes). The age at the time of death of the non-neurological controls ranged from 49 to 99 years (median: 63.5 years) with a mean post-mortem delay of 9 hours 18 minutes (SD, ± 8 hours 37 minutes). The clinical diagnosis of MS and its clinical course were determined by a certified neurologist and confirmed by a certified neuropathologist based on the neuropathological analysis of the patient’s brain autopsy.

### Neuropathological techniques and immunohistochemistry

For the classification of cortical grey matter lesions, sections were stained by immunohistochemistry for the proteolipid protein (PLP) marker of myelin. For the identification of white matter lesions, sections were stained with hematoxylin-eosin (H&E), the Luxol fast blue (LFB) marker of myelin lipids and Bielschowsky silver stain of axons. White matter lesion demyelinating and innate inflammatory activity were visualized by immunohistochemistry for PLP and the human leukocyte antigen (HLA-DR) to visualize microglia/macrophages (see antibodies details in **Supp Table 2**). Meningeal immune cells were identified by immunohistochemistry for CD3 to detect T cells, CD20 to detect B cells and Iba-1 and CD68 to detect myeloid cells (**Supp Table 2**).

Immunohistochemistry was performed as previously described (61, 62). Sections of 7µm thickness were cut from formalin-fixed paraffin-embedded tissue blocks, collected on Superfrost Plus glass slides (VWR international; Leuven, Belgium) and dried overnight at 37°C. Sections were deparaffinized in xylene (2 × 15 minutes) and rehydrated through a series (100%, 70%, 50%) of ethanol. Endogenous peroxidase activity was blocked by incubation in methanol (Merck KGaA, Darmstadt, Germany) with 0.3% H_2_O_2_ (Merck KGaA) for 20 minutes at room temperature (RT). Sections were then rinsed in PBS and pre-treated with microwave antigen retrieval (3 minutes at 900W followed by 10 minutes at 90W) in either 0.05M tris buffered saline (TBS, pH 7.6) or 10 mM Tris/1 mM ethylenediaminetetraacetic acid (EDTA) buffer pH 9.0 (**Supp Table 2**).

Sections were incubated overnight at 4°C in the appropriate primary antibody (**Supp Table 2**) diluted in Normal Antibody Diluent (Immunologic, Duiven, The Netherlands) and the next day with the BrightVision poly-HRP-Anti Ms/Rb/Rt IgG biotin-free (diluted 1:1 in PBS, ImmunoLogic) for 30 minutes at RT. The immunostaining was visualized with 3,3’-diaminobenzidinetetrahydrochloride (DAB, Vector Laboratories) for 4 minutes at RT and sections were counterstained with haematoxylin (Sigma Chemie GmbH, Steinheim, Germany), dehydrated in ethanol and mounted with Pertex (Histolab, Gothenburg, Sweden).

### Classification of cortical grey matter lesion (GML) type

Cortical GML were classified based on the location of the demyelinating plaque, as previously described (1). Leukocortical (type I) lesions were contiguous with subcortical WML; intracortical (type II) lesions were confined to the cortex and often perivascular; and subpial (type III) lesions extended from the pial surface typically to cortical layers 2-4.

### Classification of subcortical white matter lesion (WML) type

Subcortical WML are white matter lesions that lay underneath the cortical grey matter. Of note, periventricular lesions are not scored separately from other white matter lesions and are therefore included in this study as part of the assessment of subcortical WML. WML were classified as previously described (61, 63) into active, mixed active/inactive (also known as chronic active), inactive or remyelinated. Active WML typically showed loss of LFB and PLP with HLA^+^ microglia/macrophages spanning the entire lesion area. Mixed active/inactive WML typically showed loss of LFB and PLP with a hypocellular and inactive lesion center, surrounded by a rim of activated HLA^+^ microglia/macrophages (**Supp Fig 7**). Inactive WML also showed loss of LFB and PLP, though their borders were sharply demarcated with few or no HLA^+^ microglia/macrophages (**Supp Fig 8**). Remyelinated lesions are identified by thinner myelin sheaths resulting in a paler PLP^+^ staining intensity.

### White matter lesion subtype calculations

To calculate the proportions of white matter lesion subtypes, all available archived formalin-fixed paraffin-embedded (FFPE) tissue blocks for each of 27 MS patients (range 5-72 blocks, median: 30 blocks per donor) were analyzed (see **Supp Table 1**). One section per block was used and all lesions in the section that were visibly distinct were counted separately. The proportion of active lesions was defined as the number of active lesions/(the number of active + mixed active/inactive + inactive + remyelinated lesions). The proportion of mixed active/inactive lesions was defined as the number of mixed active/inactive lesions/(the number of active + mixed active/inactive + inactive + remyelinated lesions). The proportion of inactive lesions was defined as the number of inactive lesions/(the number of active + mixed active/inactive + inactive + remyelinated lesions). The proportion of remyelinated lesions was defined as the number of remyelinated lerions/(the number of active + mixed active/inactive + inactive + remyelinated lesions).

### Quantification of meningeal inflammation

Meningeal segments were randomly selected for imaging at 20x magnification with a light microscope (Olympus BX41TF, Zoeterwoude, the Netherlands) connected to the Cell D software (Olympus, Zoeterwoude, the Netherlands). Immune cells were quantified in meningeal areas that were adjacent to type III (subpial) grey matter lesions (GML) and in areas that were adjacent to normal appearing grey matter (NAGM). A total of 71±20% (mean±SD) of intact meninges were available for scoring in the MS cohort and 40±8% (mean±SD) of intact meninges were available for scoring in the NNC cohort. CD20^+^ B cell counts were done over a total meningeal area of 47.53 mm^2^ from MS patients, of which 37.02 mm^2^ was adjacent to GMLs and 10.51 mm^2^ was adjacent to NAGM; and 5.652 mm^2^ of meningeal area was adjacent to non-neurological control cortex. CD3^+^ T cell counts were done in a total meningeal area of 60.97 mm^2^ from patients, of which 48.3 mm^2^ was adjacent to GMLs and 12.67mm^2^ was adjacent to NAGM; and 6.807 mm^2^ of meningeal area was adjacent to non-neurological control cortex. Iba-1^+^ cell count was done in a total meningeal area of 46.93 mm^2^ from patients, of which 35.16 mm^2^ was adjacent to GMLs and 11.77 mm^2^ was adjacent to NAGM; and 6.66 mm^2^ of meningeal area was adjacent to non-neurological control cortex. CD68^+^ cell count was done in a total meningeal area of 44.67 mm^2^ from patients, of which 30.07 mm^2^ was adjacent to GMLs and 14.6 mm^2^ was adjacent to NAGM; and 7.97 mm^2^ of meningeal area was adjacent to non-neurological control cortex. The meningeal area (in mm^2^) was measured using the “measurement” function of the Image Pro Plus 7.0 imaging software (MediaCybernetics, Rockville, MD, USA). Cell numbers were expressed as mean number per mm of intact meninges.

### Statistics

All tests were performed using GraphPad Prism software 5.0 (GraphPad Software Inc, San Diego, CA, USA). The variability of distribution was assessed by Shapiro-Wilk normality test. The non-normally distributed data was analysed by the non-parametric Mann Whitney test (between two groups) or Kruskal-Wallis test followed by Dunn’s correction for multiple comparisons (between more than two groups). Results were considered significant when p <0.05 at a 95% confidence interval.

### Study approval

All post-mortem human tissue was collected with informed consent for the use of material and clinical data for research purposes, in compliance with ethical guidelines of the Vrij Universiteit and Netherlands Brain Bank, Amsterdam, The Netherlands (Reference 2009/148). In addition the University of Toronto Research Ethics Board (REB) granted approval for conducting histology and correlation with clinical metadata on all post-mortem human tissue (study number: 36850).

## Data Availability

All data produced in the present study are available upon reasonable request to the authors

## Author’s contribution

SMA conducted the experiments and analyzed the data; NF acquired data; HT, JLG and IH advised on the project; IM and VR participated in the initial data acquisition; ABO and VR designed the study; VR wrote the manuscript.

## Acknowledgements

This study was funded by the National Multiple Sclerosis Society (grant RG 4775A1/1). We thank the donors of the Netherlands Brain Bank.

## Notes

### Competing Interest Statement

The authors have declared no competing interest.

