## Supplemental material for "Accumulation of meningeal lymphocytes, but not myeloid cells, correlates with subpial cortical demyelination and white matter lesion activity in progressive MS patients"

***Corresponding authors:**

**Supplemental Figures:**

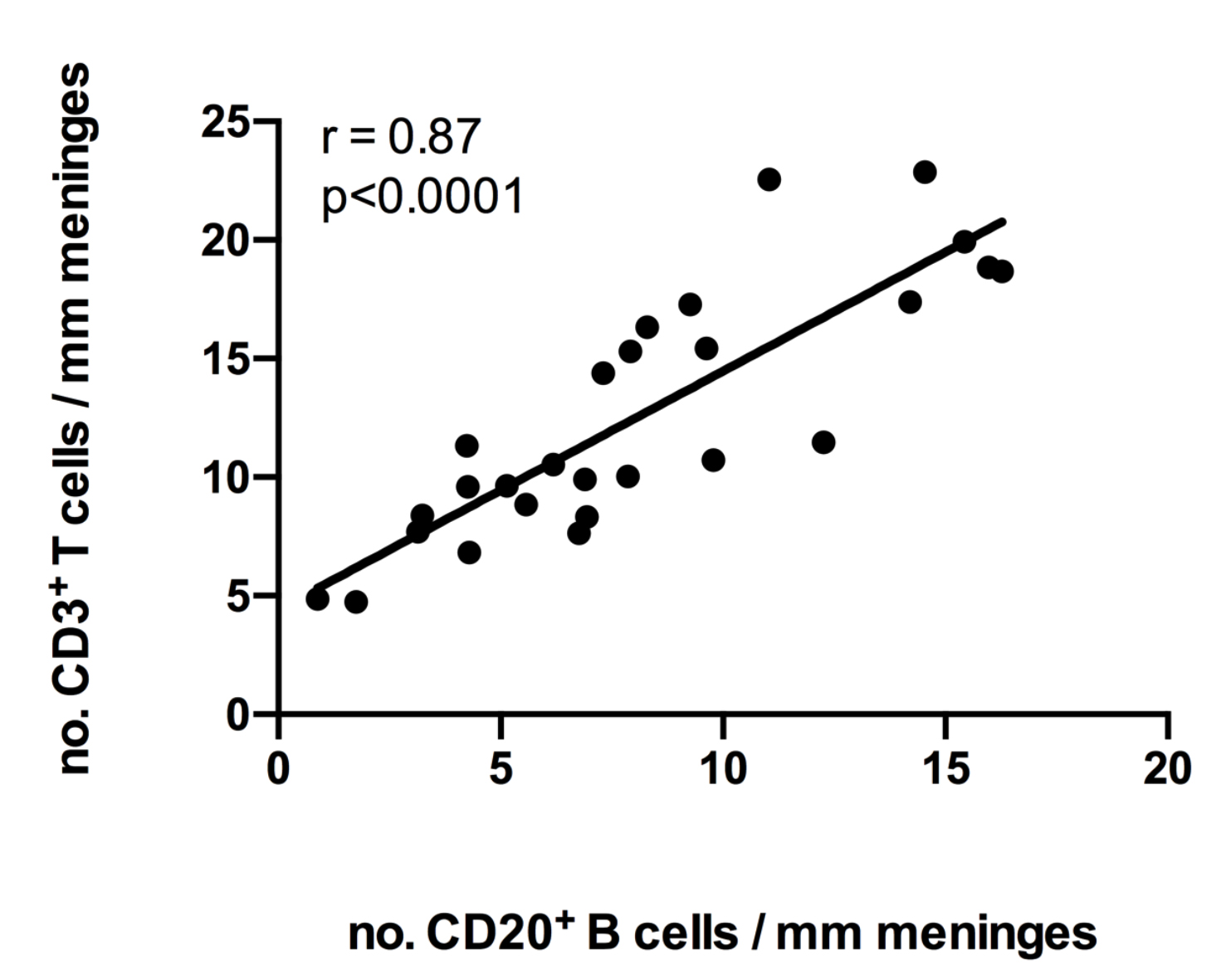

**Supplementary figure 1. The density of meningeal T cells positively correlates with the density of meningeal B cells in MS.** Spearman correlation coefficient between meningeal CD3^+^ T cell count and meningeal CD20^+^ B cell count of MS donors. Each data point represents the mean meningeal cell count in all fields analyzed per case.

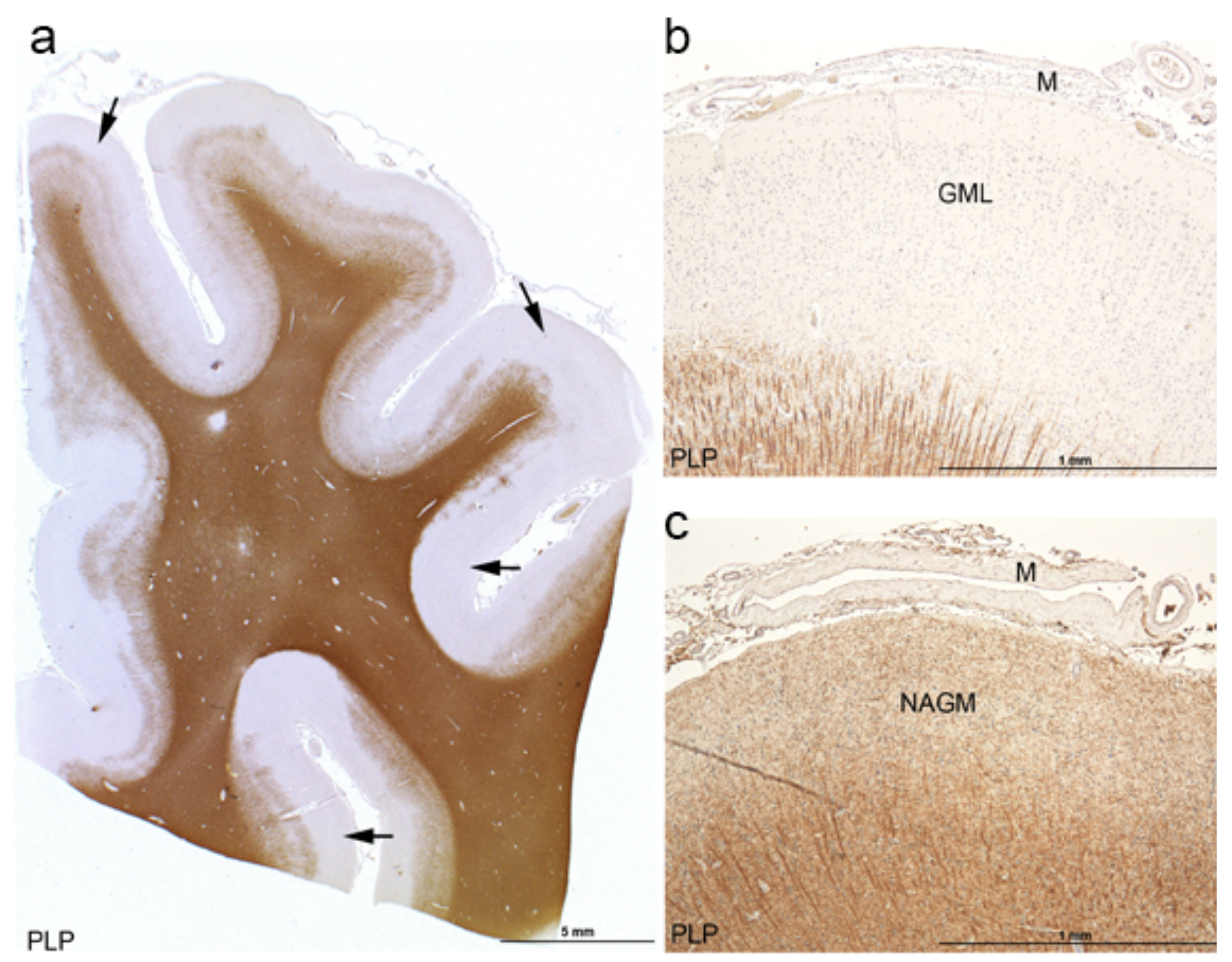

**Supplementary figure 2. Cortical subpial demyelination in MS. a-c**. Immunohistochemical staining with anti-myelin proteolipid protein (PLP) of MS cortex with subpial grey matter demyelination (arrows in **a**). Close up of MS cortex showing a subpial (type III) grey matter lesion (GML) (**b**) or normal appearing grey matter (NAGM) (**c**) adjacent to meninges (M). In **a**, scale bar represents 5mm; In **b**, **c**, scale bars represent 1mm.

**Supplementary figure 3. Meningeal T cells and B cells in the ‘low’ donor group are not enriched in proximity of cortical subpial demyelinated lesions.** Representative immunohistochemical staining for CD3 (**a and b, arrows**) and CD20 (**c and d, arrows**) in meninges adjacent to a subpial grey matter lesion (GML) or adjacent to normal appearing gray matter (NAGM) in MS donors with low CD3^+^ or CD20^+^ meningeal cell count. **e., f.** Representative immunostaining for CD3 and CD20 in Lymph Nodes used as technical positive controls. In **a-d**, scare bars represent 100μm. In **e, f**, scare bars represent 200μm.

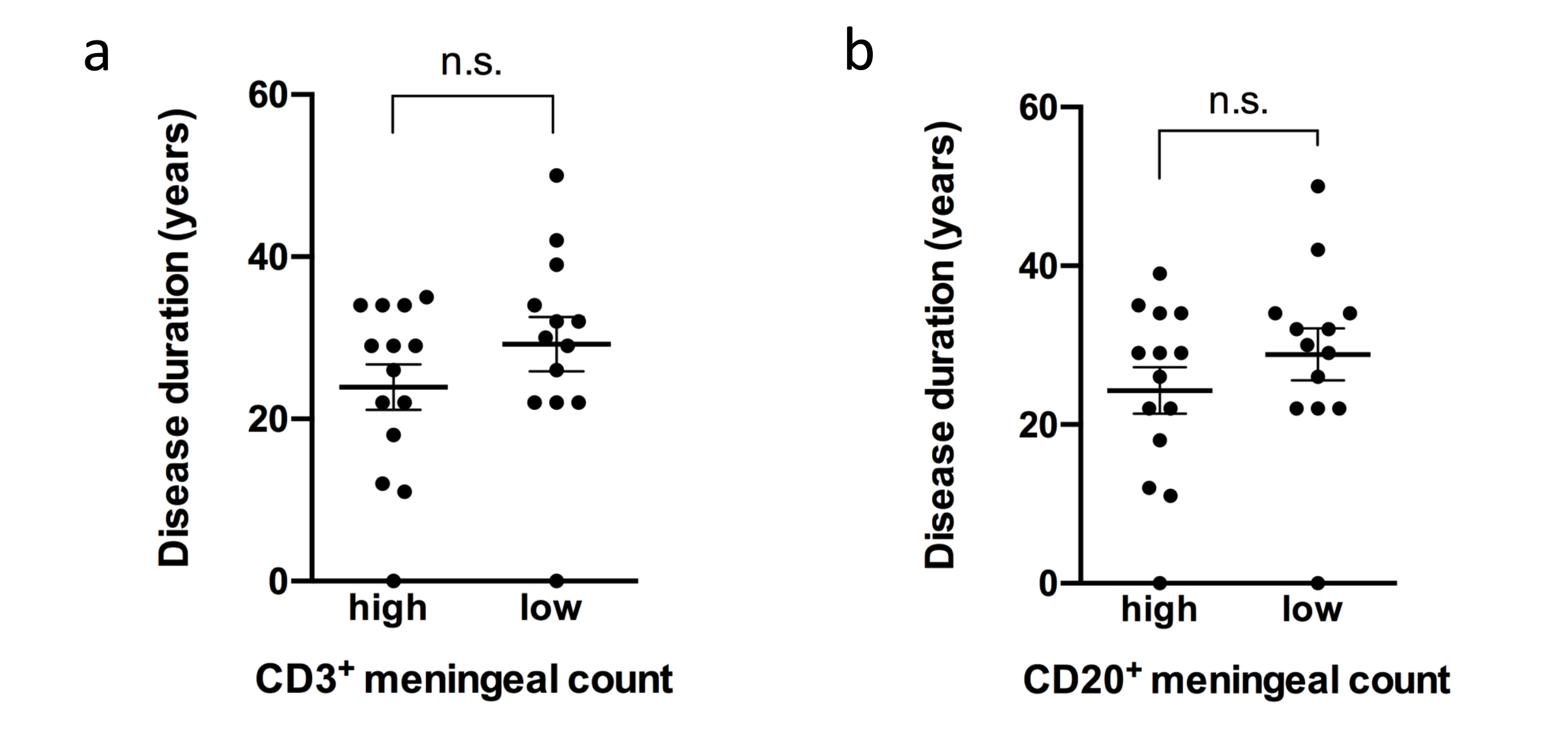

**Supplementary figure 4. Enrichment of meningeal T cells and B cells is not linked to disease duration.** Disease duration in MS donors with high vs low (**a**) CD3^+^ or (**b**) CD20^+^ meningeal cell count. Statistically significant differences were tested by the non-parametric Mann Whitney test (p<0.05).

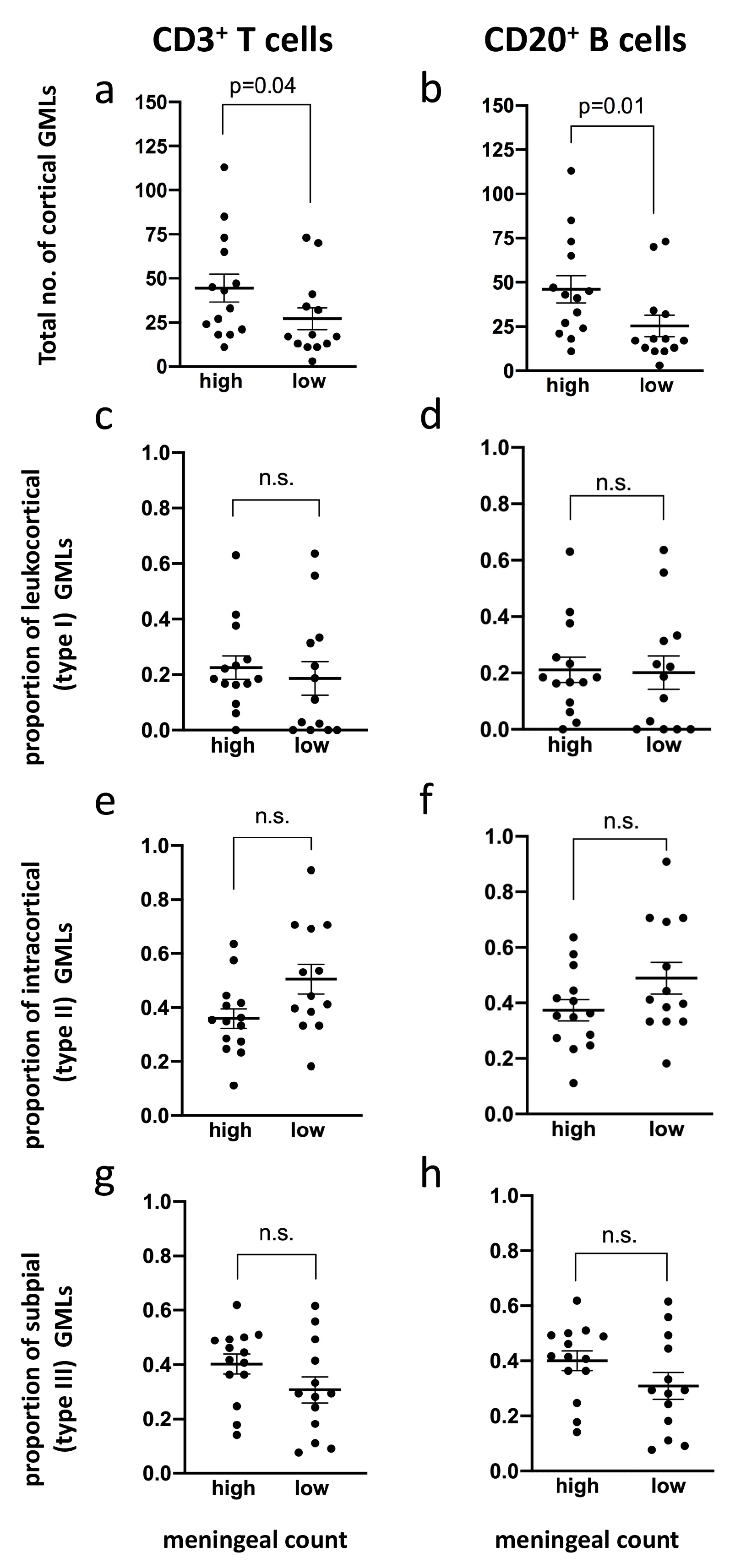

**Supplementary figure 5. Enrichment of meningeal T cells and B cells is not linked to the proportion of cortical subpial demyelinated grey matter lesions.** Quantification of (**a, b**) proportion of cortical grey matter lesions (GMLs), (**c, d**) proportion of leukocortical (type I) GMLs, (**e, f**) proportion of intracortical (type II) GMLs and (**g, h**) proportion of subpial (type III) GMLs in MS donors with high vs low meningeal CD3^+^ T cells (**a, c, e, g**) or CD20^+^ B cell (**b, d, f, h**) count. Each data point represents the proportion of GMLs in all tissue blocks analyzed per case. Statistically significant differences were determined by the non-parametric Mann Whitney test

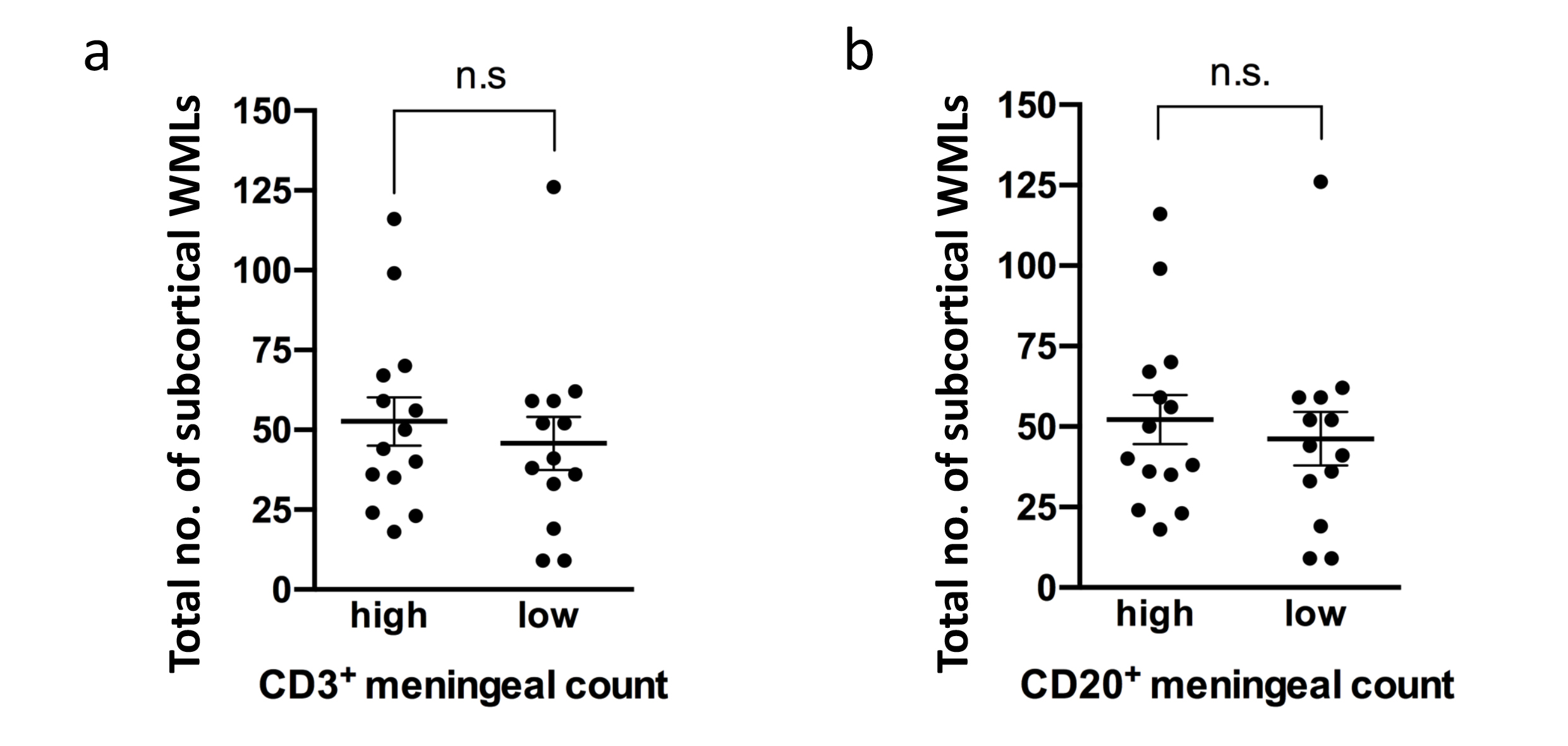

**Supplementary figure 6. Enrichment of meningeal T cells and B cells is not linked to the total number of subcortical white matter lesions.** Total number of subcortical white matter lesions in MS donors with high vs low (**a**) CD3^+^ or (**b**) CD20^+^ meningeal cell count. Statistically significant differences were tested by the non-parametric Mann Whitney test (p<0.05).

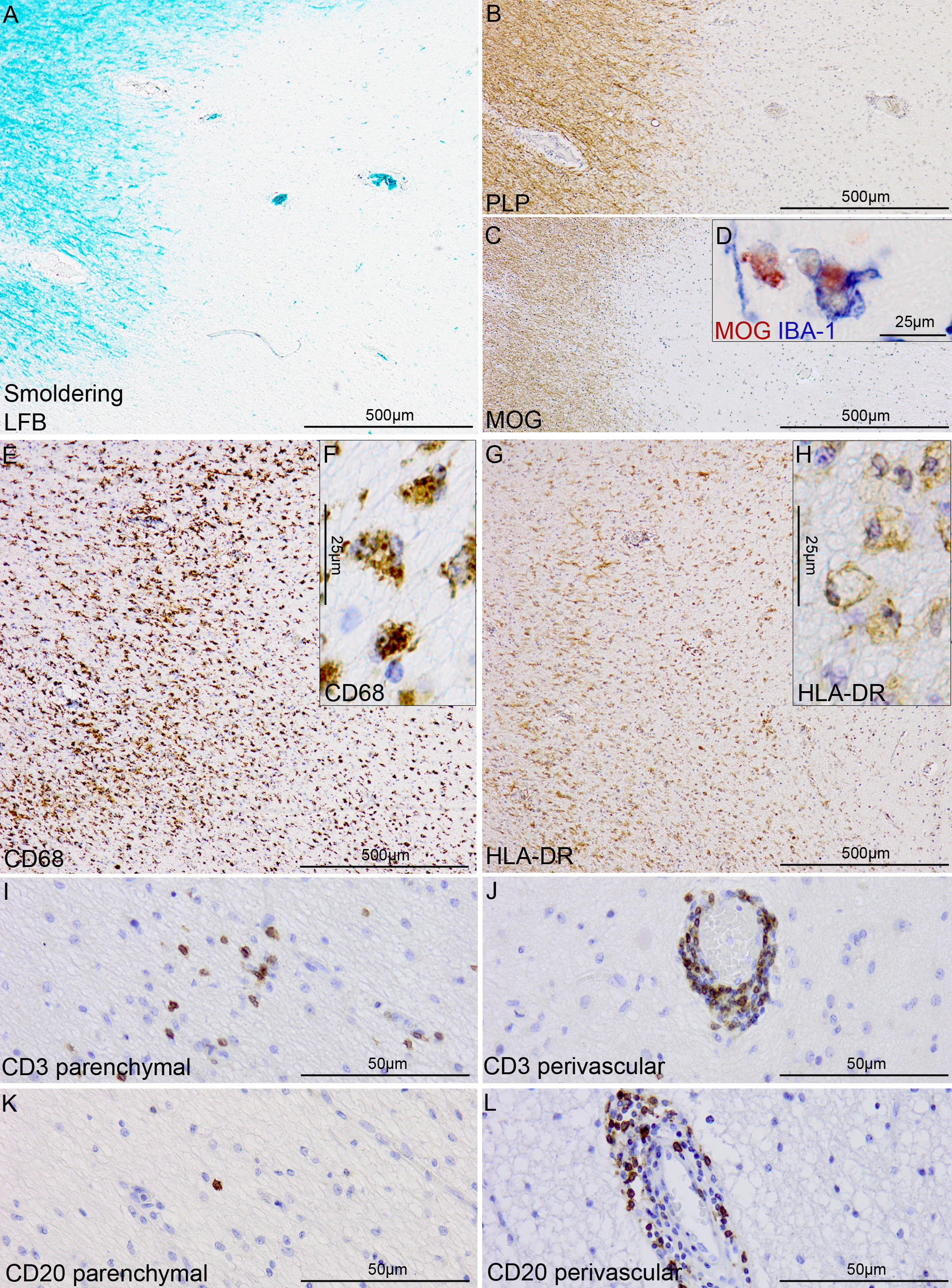

**Supplementary figure 7. Characterization of mixed active-inactive white matter lesions (WLM) in progressive MS cases.** A mixed active-inactive WML in progressive MS case, showing loss of luxol fast blue (LFB) staining (**a**), loss of immunoreactivity for the proteolipid protein (PLP, in **b**) and loss of immunoreactivity for the myelin oligodendrocyte glycoprotein (MOG, in **c**) within the lesions, with intracellular inclusions of MOG+ myelin products inside ionized calcium-binding adapter molecule-1 (IBA-1)+ macrophages at the lesion edge (**d**), indicative of demyelinating activity. Immunoreactivity for the CD68 lysosomal marker is abundant throughout the lesion and accumulates at the lesion edge (**e**) in a staining pattern consistent with enlarged lysosomes within phagocytic cells (**f**). Human leukocyte antigen (HLA-DR)+ myeloid cells accumulate at the lesion edge (**g**) and show enlarged morphology (**h**), consistent with the active phenotype. CD3+ T cells are found in the parenchyma (**i**) and as perivascular “cuffs”, expanding within the Virchow-Robbin space of capillaries (**j**), located within the lesion or at the peri-lesional areas. CD20+ B cells are occasionally found in the parenchyma (**k**) and more often observed as perivascular “cuffs” (**l**), located within the lesion or at the peri-lesional areas.

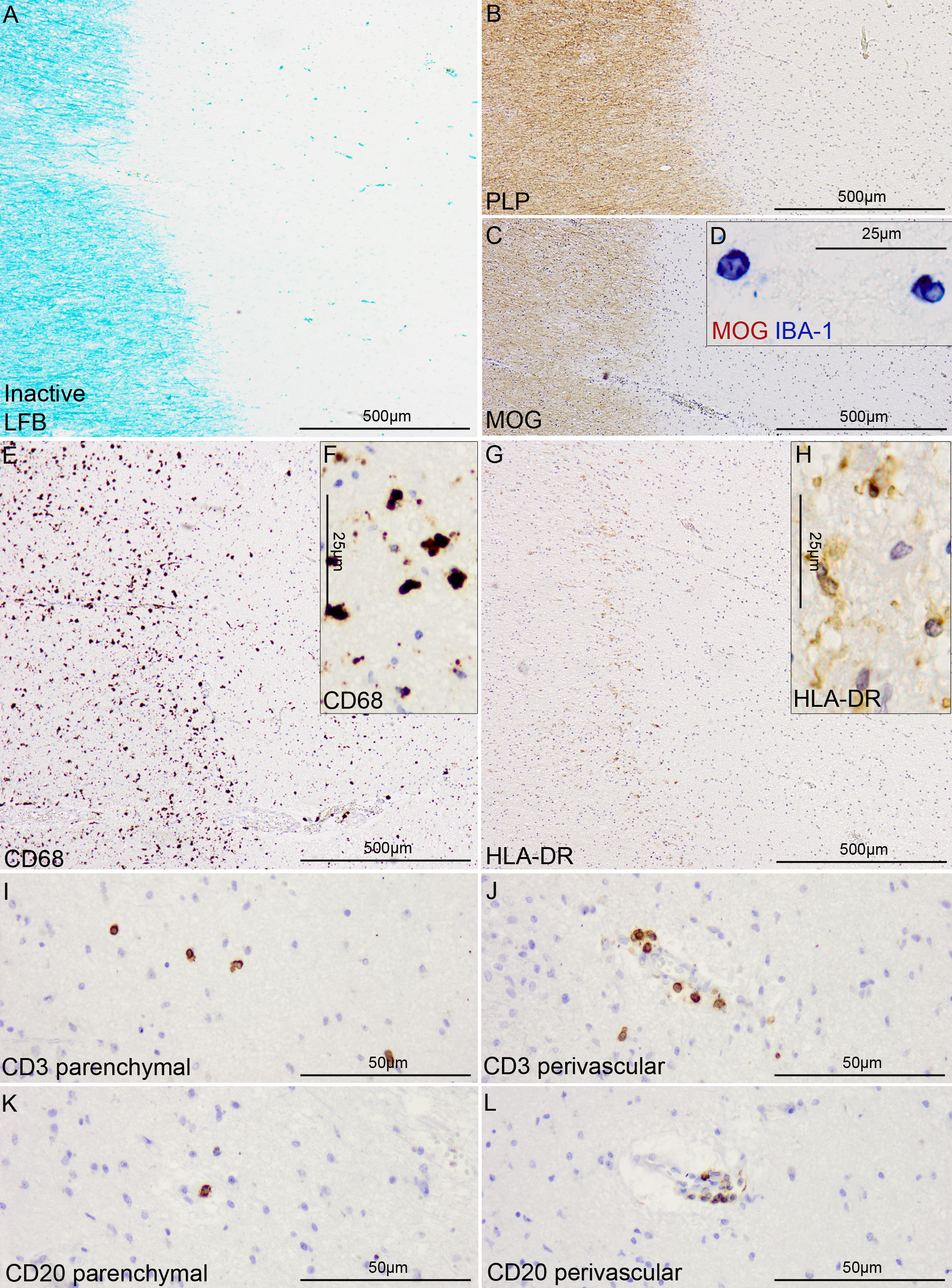

**Supplementary figure 8. Characterization of inactive WML in progressive MS cases.** Inactive WML in a progressive MS case, showing loss of luxol fast blue (LFB) staining (**a**), loss of immunoreactivity for the proteolipid protein (PLP, in **b**) and loss of immunoreactivity for the myelin oligodendrocyte glycoprotein (MOG, in **c**) with no evidence of intracellular inclusions of MOG+ myelin products inside ionized calcium-binding adapter molecule-1 (IBA-1)+ macrophages (**d**), indicative of no demyelinating activity. Immunoreactivity for the CD68 lysosomal marker is scarce within the lesion and more obvious at the lesion edge (**e**) in a staining pattern consistent with inactive, gliotic cells (**f**). A paucity of human leukocyte antigen (HLA-DR)+ microglia can be found at the lesion edge (**g**) and show small ramified morphology (**h**), consistent with the inactive, resting phenotype. (**i-l**) CD3+ T cells and CD20+ B cells are occasionally found in the parenchyma (**i, k**) and perivascular space (**j, l**), located within the lesion or at the peri-lesional area.

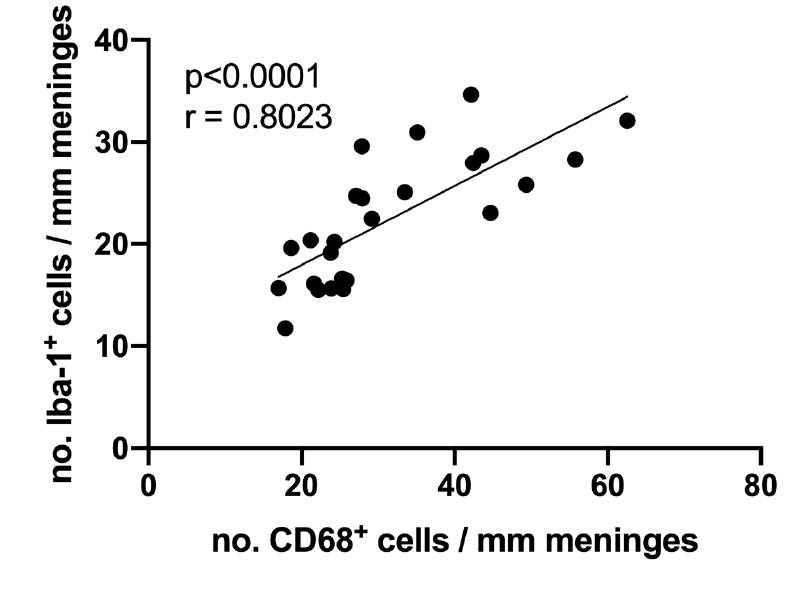

**Supplementary figure 9. The density of meningeal Iba-1^+^ myeloid cells positively correlates with the density of meningeal CD68^+^ cells in MS.** Spearman correlation coefficient between meningeal Iba-1^+^ myeloid cell count and meningeal CD68^+^ cell count of MS donors. Each data point represents the mean meningeal cell count in all fields analyzed per case.

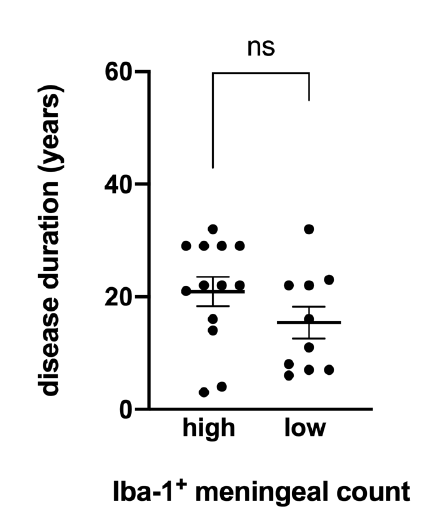

**Supplementary figure 10. Enrichment of meningeal myeloid cells is not linked to disease duration.** Disease duration in MS donors with high vs low Iba-1^+^ meningeal cell count. Statistically significant differences were determined by the non-parametric Mann Whitney test (p<0.05).

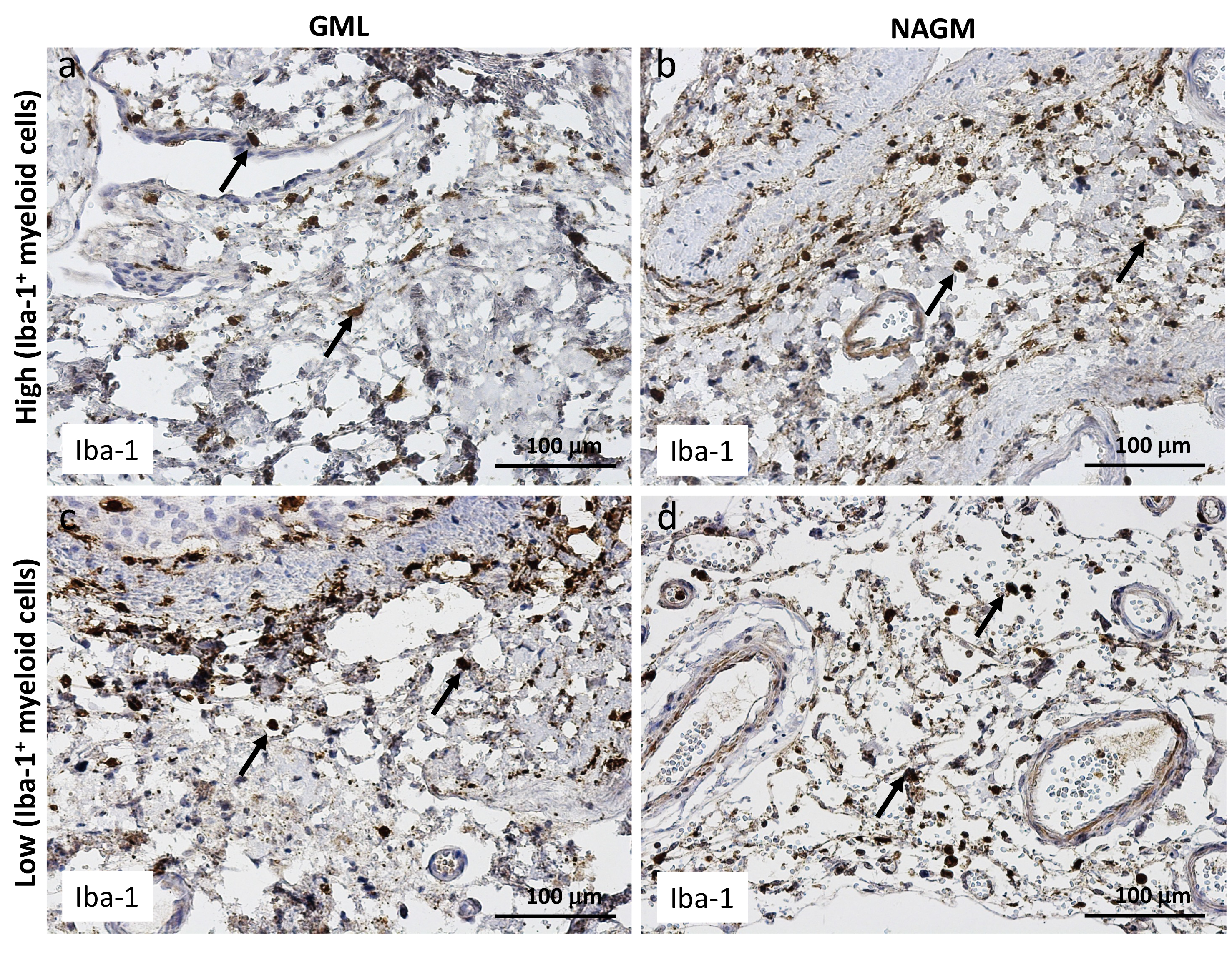

**Supplementary figure 11. Meningeal myeloid cells are not topographically linked to subpial demyelination in MS.** Representative immunohistochemical staining for Iba-1 (arrows) in meninges adjacent to a subpial grey matter lesion (GML) or adjacent to normal appearing gray matter (NAGM) in MS donors with high (**a, b**) or low (**c, d**) meningeal Iba-1^+^ myeloid cell count. In **a-d**, scare bars represent 100μm.

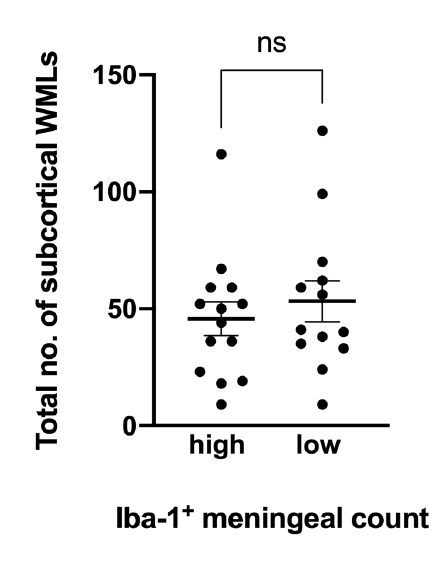

**Supplementary figure 12. Enrichment of meningeal myeloid cells is not linked to the total number of subcortical white matter lesions.** Total number of subcortical white matter lesions in MS donors with high vs low Iba-1^+^ meningeal cell count. Statistically significant differences were determined by the non-parametric Mann Whitney test (p<0.05).

**Supplemental Tables:**

| **Supplementary table 1. Donor demographics** | | | | | | | |
| --- | --- | --- | --- | --- | --- | --- | --- |
| **Case** | **Sex** | **Age range**  **(years)** | **PMD**  **(h:min)** | **Type of MS** | **DD**  **(years)** | **No. tissue blocks analysed for lesion characterization** | **COD** |
| ***MS*** | | | | | | | |
| 1 | F | 48-81 | 08:40 | SPMS | 26 | 41 | Respiratory insufficiency to (uro)sepsis |
| 2 | F | 48-81 | 10:40 | SPMS | 29 | 30 | Euthanasia |
| 3 | F | 48-81 | 07:30 | SPMS | 34 | 27 | Euthanasia |
| 4 | F | 48-81 | 11:50 | PPMS | 22 | 32 | Respiratory failure with end stage MS |
| 5 | M | 48-81 | 08:15 | SPMS | 30 | 23 | Pneumonia, cachexia and dehydration |
| 6 | F | 48-81 | 09:05 | SPMS | 18 | 23 | Euthanasia |
| 7 | F | 48-81 | 08:35 | PPMS | 29 | 34 | Aspiration pneumonia |
| 8 | F | 48-81 | 05:45 | PPMS | 22 | 15 | Sepsis |
| 9 | F | 48-81 | 08:25 | SPMS | 11 | 8 | Natural death |
| 10 | M | 48-81 | 11:00 | SPMS | >12 | 38 | Exact cause unknown, infection 2 days prior to death |
| 11 | F | 48-81 | 08:40 | PPMS | 29 | 31 | Euthanasia |
| 12 | F | 48-81 | 08:25 | SPMS | 34 | 27 | Respiratory insufficiency secondary to pneumonia |
| 13 | M | 48-81 | 07:30 | - | - | 28 | - |
| 14 | F | 48-81 | 06:45 | PPMS | 29 | 5 | Cardiac asthma |
| 15 | F | 48-81 | 07:30 | SPMS | 39 | 23 | Bronchitis/ aspiration pneumonia |
| 16 | M | 48-81 | 10:45 | RRMS | 22 | 34 | Euthanasia |
| 17 | F | 48-81 | 08:00 | SPMS | 42 | 32 | Pneumonia |
| 18 | M | 48-81 | 06:20 | PPMS | 32 | 33 | Respiratory insufficiency |
| 19 | F | 48-81 | 10:05 | SPMS | 26 | 18 | Cardiovascular event and dehydration |
| 20 | F | 48-81 | 07:50 | SPMS | 50 | 72 | Euthanasia |
| 21 | M | 48-81 | 10:45 | RRMS | 22 | 34 | Euthanasia |
| 22 | F | 48-81 | 07:05 | SPMS | 34 | 26 | Cachexia with slowly progressive MS and metastatic breast cancer |
| 23 | F | 48-81 | 09:35 | PPMS | 22 | 25 | Cardiac asthma |
| 24 | F | 48-81 | 04:35 | SPMS | 34 | 30 | Aspiration pneumonia |
| 25 | M | 48-81 | 09:15 | - | - | 30 | - |
| 26 | M | 48-81 | 07:30 | PPMS | 32 | 30 | Respiratory failure due to pneumonia |
| 27 | F | 48-81 | 09:45 | SPMS | 35 | 34 | Euthanasia |
| ***Non-neurological Controls*** | | | | | | | |
| 28 | M | 49-99 | 06:15 | - | - | - | Euthanasia with Hopkin’s lymphoma |
| 29 | F | 49-99 | 04:15 | - | - | - | - |
| 30 | M | 49-99 | 26:45 | - | - | - | Pulmonary infection |
| 31 | F | 49-99 | 07:30 | - | - | - | Infection e.c.i |
| 32 | M | 49-99 | 05:00 | - | - | - | - |
| 33 | F | 49-99 | 06:00 | - | - | - | Cachexia |
| 34 | M | 49-99 | 14:00 | - | - | - | Heart failure |
| 35 | F | 49-99 | 7:40 | - | - | - | - |
| 36 | M | 49-99 | 7:44 | - | - | - | - |
| MS,multiple sclerosis; F, Female; M, Male; PMD, post-mortem delay; DD, disease duration; h:min, hours:minutes; COD, cause of death. | | | | | | | |

| **Supplementary table 2. Primary antibodies, dilution, source** | | | |
| --- | --- | --- | --- |
| Antigen | Clone | Dilution/Concentration | Source |
| Proteolipid protein (PLP) | Monoclonal (plpc1) | 1:3000^a^ | Serotec (Puchheim, Germany) |
| CD3 | Monoclonal (SP7) | 1:500^b^ | Dako (Glostrup, Denmark) |
| CD20 | Monoclonal (L26) | 1:1000^b^ | Dako |
| Iba-1 | Monoclonal (EPR16589) | 1:8000^c^ | Abcam |
| CD68 | Monoclonal (PGM1) | 1:200^b^ | Dako |
| Human leukocyte antigen (HLA)-DR | Monoclonal (CR3/43) | 8.6μg/ml^a^ | Dako |
| Antigen retrieval of paraffin sections was performed by heat in ^a^ 0.05 M Tris buffered saline pH 7.6; ^b^ 10 mM Tris/1 mM EDTA buffer pH 9 or 10mM Sodium Citrate pH 6.0^c^ | | | |
